# Nature-based social prescribing programmes: opportunities, challenges, and facilitators for implementation

**DOI:** 10.1101/2023.11.27.23299057

**Authors:** Siân de Bell, Julius Cesar Alejandre, Claudia Menzel, Rita Sousa-Silva, Tanja M. Straka, Susanne Berzborn, Max Bürck-Gemassmer, Martin Dallimer, Chris Dayson, Jessica C. Fisher, Annette Haywood, Alina Herrmann, Gisela Immich, Christian S. Keßler, Kristin Köhler, Mary Lynch, Viola Marx, Andreas Michalsen, Pierpaolo Mudu, Hendrik Napierala, Maximilian Nawrath, Sharon Pfleger, Claudia Quitmann, Jonathan P. Reeves, Kevin Rozario, Wolfgang Straff, Katie Walter, Charlotte Wendelboe-Nelson, Melissa R. Marselle, Rachel Rui Ying Oh, Aletta Bonn

## Abstract

**Background:** Evidence on the health benefits of spending time in nature has highlighted the importance of provision of blue and green spaces in people’s living environments. The potential for health benefits offered by nature exposure, however, extends beyond health promotion to health treatment. Social prescribing links people with health or social care needs to community-based, non-clinical health and social care interventions. The aim is to improve health and wellbeing. Nature-based social prescribing (NBSP) is a variant which uses the health-promoting benefits of activities carried out in natural environments, such as gardening and conservation volunteering. Much of current NBSP practice has been developed in the UK and there is increasing global interest in its implementation. This requires interventions to be adapted for different contexts, considering the needs of populations and the structure of healthcare systems.

**Methods:** This paper presents results from an expert group participatory workshop involving 29 practitioners, researchers, and policymakers from the UK and Germany’s health and environmental sectors. Using the UK and Germany, two countries with different healthcare systems and in different developmental stages of NBSP practice as case studies, we analysed opportunities, challenges, and facilitators for the development and implementation of NBSP.

**Results:** We identified five overarching themes for developing, implementing, and evaluating NBSP: *Capacity Building*; *Universal Accessibility*; *Embedded and Integrated Networks and Collaborations*; *Standardised Implementation and Evaluation*; and *Sustainability*. We also discuss key strengths, weaknesses, opportunities, and threats (i.e., a SWOT analysis) for each overarching theme to understand how they could be developed to support NBSP implementation.

**Conclusions:** NBSP could offer significant public health benefits using available blue and green spaces. We offer guidance on how NBSP implementation, from wider policy support to the design and evaluation of individual programmes, could be adapted to different contexts. This research could help inform the development and evaluation of NBSP programmes to support planetary health from local and global scales.

## Introduction

Currently, healthcare systems across the world tend to focus on the treatment of diseases rather than health promotion or prevention. The use of pharmaceuticals is the most common intervention in healthcare (1) and as a result, a large proportion of national healthcare budgets are allocated to pharmaceutical costs. The pharmaceutical industry is continuously growing, with the global market dominated by countries from North America (USA and Canada) and Europe (2). In high-income countries like Germany and the UK, pharmaceuticals make up a large proportion of total health spending, 13.87% and 9.46% respectively in 2021 (3).

With increased spending on pharmaceutical interventions to treat ill-health, their effectiveness, costs, and impacts on the climate, environment, and biodiversity are of rising concern (4,5). For example, the carbon emissions of the pharmaceutical industry are more than that of the automotive industry (6), and in the UK, pharmaceutical acquisition accounts for around 25% of National Health Service (NHS) carbon emissions (7). Furthermore, the use of pharmaceuticals correlates with the concentration of active pharmaceutical ingredients in terrestrial and aquatic environments, affecting water quality, soil health, food crops, and the lives of various species (5,8). For these reasons, non-pharmaceutical health interventions, treatments, and health and wellbeing measures are needed to help reduce the pressures of pharmaceutical consumption on planetary health (Box 1)(9).

### Social prescribing – an alternative to pharmaceutical interventions

Social prescribing is one alternative to pharmaceutical treatments and may help to reduce their use. Activities that could be considered as social prescribing have been used for many years to support health and wellbeing, with language and formal systems being developed more recently, particularly in the UK (10). While there is work in progress to establish a common definition for social prescribing (11), it is usually considered to be a process by which people with health and/or social care needs are connected with community-based services and activities. These are typically provided by the local voluntary and community sector, and primarily address the social determinants of health (Box 1). There is growing global interest in social prescribing, with its implementation in a range of countries, and support emerging at the international level (11,12). For example, the Global Social Prescribing Alliance, an independent group of international partners from over 20 countries, was formed in 2021 to facilitate knowledge exchange and to share ideas and best practice on social prescribing (13). Additionally, the WHO Regional Office for the Western Pacific has published a training toolkit to support the implementation of social prescribing (14).

Typically, the process of social prescribing involves an ‘identifier’, who recognises a person with a health or social care need (referred to as a service user or patient), and a ‘connector’ (who may also be the identifier), who refers them to a community-based service or activity with the aim of improving their health and wellbeing (11,15). These identifiers and connectors differ between healthcare systems (12). Identifiers might work in a clinical setting (e.g., as a primary care physician) or in the community. In a direct referral pathway, they are also the connector, or they may refer the service user to a connector (e.g., a link worker pathway) (11,15) (Box 1). The connector and service user explore the service user’s priorities and needs to match them with a compatible activity or service (11,16); these could range from information, advice or support services, to physical, arts, or nature-based activities (17).

Nature-based social prescribing (Box 1) is a specific form of social prescribing, which involves a person being referred to an activity taking place in nature (18,19). These activities may include active connection with nature, such as walking or gardening, or a passive one (e.g., receiving a talking therapy outdoors). The activities aim to utilise the health benefits that have been found to be linked with spending time in nature (see below). Co-benefits could include restoring and maintaining ecosystems and biodiversity (e.g., conservation-based programmes), increasing people’s connection with nature, and their social connections, and the development or improvement of pro-environmental behaviours (20).

##### Box 1 Definitions related to nature-based social prescribing

- **Nature** is defined as physical features and non-human processes (e.g., weather), including “living nature” (plants and animals), and the landscapes (e.g., forest, river, lake, urban green park) that comprise these (21). These landscapes, or natural environments, might also be referred to as green spaces, or where they include water (e.g. rivers, lakes, canals), blue spaces (22).
- **Direct nature contact** is being physically present in a natural environment, while **indirect nature contact** is experiencing nature without being physically present in it (e.g., window view, virtual nature) (23).
- **Planetary health** is “the achievement of the highest attainable standard of health, wellbeing, and equity worldwide through judicious attention to the human systems— political, economic, and social—that shape the future of humanity and the Earth’s natural systems that define the safe environmental limits within which humanity can flourish” (p. 1978, (24).)
- **Prescribing** is a formal clinical or pharmacological intervention, usually evidence-based and provided by a trained and licensed healthcare provider (i.e., identifier; e.g., doctor, psychotherapist, pharmacist, nurse prescriber) to patients with the aim of treating or preventing an illness, according to clinical guidelines, national rules, and legislation.
- **Social prescribing** is a process through which service users with an identified health and/or social care need are connected from a healthcare or community setting to non-clinical services, with the aim of improving their health and wellbeing and strengthening their community connections. This social prescription is co-produced by the service user and the person who connects them with the non-clinical service (11,25). Non-clinical services are commonly delivered by voluntary sector or community-based organisations and include a range of activities (e.g. arts, physical activity, information/advice/support, and nature) (17).
- **Link worker (social prescribing) pathway** employs the services of a **link worker** (i.e., a **connector**, sometimes known by other names such as ‘community navigators’), a care professional usually employed in a primary care setting. They spend time with service users who are usually referred by a member of the primary healthcare team (i.e. the identifier, e.g., general practitioners [GPs], nurses) to identify their needs and preferences to make appropriate referrals to community activities (12,15,26).
- **Nature-based health intervention (NBHI)** are programmes or strategies that engage people with nature to improve their health and wellbeing. NBHI might include changes to the natural environments where people live and work, or activities that take place in nature, such as therapeutic fly-fishing or forest bathing (27).
- **Nature-based social prescribing (NBSP) (or nature prescription, green prescription, blue prescription)** is a type of social prescribing and a type of NBHI. Service users are prescribed to activities that are carried out in nature, utilising the health-promoting benefits of time spent in nature. Activities might include walks, gardening, surfing, or simply spending time in natural environments (18,28).

### The benefits of time spent in nature

Numerous mechanisms have been proposed for how nature might affect health and wellbeing (for a recent analysis of data from 18 countries, see (29)). For instance, nature can reduce harm (e.g., decreased exposure to pollution), restore capacities (e.g., enable recovery from stress), and build capacities (e.g., facilitate physical activity) (23). However, nature can also cause harm by, for example, increasing the likelihood of infections from human exposure to vector-borne diseases, pollen allergens, or trees trapping air pollution within streets (23). The diversity of mechanisms is mirrored by the myriad ways in which people experience and relate to nature, something that is often influenced by their own experiences, memories, cultures, and local contexts (30,31).

Whilst research unpicking these mechanisms, and other factors influencing the relationship between nature and health, is still ongoing, research on the outcomes of time spent in nature has shown associations with a multitude of physical, mental, and social health and wellbeing benefits (32,33). These include improved mental health and wellbeing, positive effects on cardiovascular, respiratory and metabolic outcomes, sleep, and immunity (33,34). Moreover, exposure to nature has been demonstrated to be protective against risk factors for mental ill-health such as psychological stress (35–37).

Nature’s contribution to public health is increasingly being recognised (38–40), with consideration of the types and characteristics of green spaces, and the spatial provision and maintenance of natural environments (41,42). These environmental set-ups are important both close to people’s homes and elsewhere for recreational visits (19,23,43). However, the incorporation of proactive nature-based health interventions (NBHI, Box 1), such as NBSP, into healthcare systems as a type of treatment is not (yet) widespread.

### Current implementation of nature-based social prescribing programmes

Currently, there is no common definition for NBSP, but universal definitions and terms in the respective local language are essential both for commissioning and providing NBSP, as well as for communication with service users. There are some examples of the use of NBSP under different names. Some of the earliest studies come from New Zealand, where the Ministry of Health developed ‘green prescriptions’ in 1997 to increase physical activity outdoors (44). Similarly in the United States, there are over 75 ‘park prescription’ programmes, in which a healthcare provider gives patients a written recommendation to do physical activity in parks (45). In Germany, treatments at a spa or a sanatorium, such as Kneipp therapy or climatotherapy, can be considered NBHIs (46). In some cases, they are reimbursable by statutory health insurance as part of rehabilitation or prevention programmes; they are, however, part of a distinct tradition in healthcare (health resort medicine). Although social prescribing is not a formalised practice in Germany (12), Germany’s first social prescription service was established in 2017 (47). With increasing interest in the development of NBSP and wider social prescribing practice, Germany offers a case study for a model of NBSP in an early developmental stage.

The UK, in contrast, was the first country to integrate social prescribing into its national health policy (10), with many of its social prescribing programmes involving nature-based activities such as gardening, walking, open-water swimming, or nature conservation (18,19,23). While the initial development of NBSP was bottom-up, there is now top-down investment happening across the UK. In England, the UK Government has invested in the expansion of NBSP through a cross-sectoral initiative with a range of national partners including the Department for Environment, Food and Rural Affairs (Defra), NHS England, Sport England, and the National Academy for Social Prescribing (16). Similar investment has been made by the Scottish Government to pilot Green Health Partnerships in four Scottish areas (e.g., Ayrshire, Dundee, Highland, and Lanarkshire) with continuing expansion in other local authorities through coordination between national partners - NatureScot, Scottish Forestry, Public Health Scotland, Transport Scotland - and local partners (48).

Specific examples from the UK, operating at the local level, include Green Health Prescriptions which were developed in Scotland under the Green Health Partnership (49). These aim to reduce obesity and promote mental and physical health using local green spaces and connecting with nature (Lafferty and Finton, 2015). In Dundee, for instance, Green Health Prescriptions allow healthcare professionals in primary and secondary care to refer service users to a green health worker. Based on their interests and health goals, the service user can then access one of over 60 local nature-based community activities (Marx & More, 2022). Blue spaces (Box 1) are also being used for NBSP, with activities including surfing, kayaking, and canoeing (25). In England, the Blue Prescribing Project, which is run by the Wildfowl & Wetlands Trust (WWT) and The Mental Health Foundation, provides structured wetland experiences for individuals experiencing mental health problems (50–52). As can be seen, there are a variety of NBSP programmes in the UK, with a mix of top-down and bottom-up support for implementation. The UK therefore provides a case study for a possible advanced model of NBSP development and implementation, with ongoing evaluation through the UK Government’s Green Social Prescribing test and learn pilots monitoring its effectiveness and further supporting the use of NBSP in the healthcare system (48).

### Aims and objectives

Despite increasing global interest, successful implementation of NBSP in different countries requires adaption to fit different contexts (15,53), considering the demographics and needs of the population, the local health and social care systems, and the accessibility and suitability of natural environments. In this paper, we address opportunities, challenges, and facilitators for the development and implementation of NBSP. We focus on two countries with different NBSP models: the UK, which has a relatively advanced model of NBSP, and Germany, which represents an initial development stage.

We present perspectives from a trans- and interdisciplinary expert group workshop of health and environmental practitioners, researchers, and policymakers, mainly from the UK and Germany. The workshop allowed knowledge exchange, sharing of best practices in NBSP, and exploration of the potential for its development and implementation internationally. We aimed to answer three questions:

1. What are the opportunities for NBSP and likely health and wellbeing outcomes?
2. What are the challenges to implementing NBSP programmes?
3. What is needed to implement a pragmatic NBSP programme?

We synthesise the findings and provide recommendations that could inform healthcare providers, social prescribing practitioners, policymakers, and other stakeholders in developing and evaluating NBSP programmes in different contexts (e.g., healthcare settings and systems, countries, regions, or cities) to support planetary health at local and global scales.

## Methods

To understand the current landscape of NBSP and to identify opportunities, challenges, and ways forward for implementation, we held a two-day interdisciplinary and participatory expert group workshop in November 2022 in Berlin, Germany.

### Participant selection

The workshop was organised by a team of 10 researchers in the fields of public health, environmental psychology, and ecology. It was attended by 29 experts (excluding the organisers), from both the health and environmental sectors, with representation of research, policy, and practice organisations in the UK, Germany, and other European countries (Table 1). Workshop participants were invited by identifying relevant organisations and representatives through professional networks and targeted online searches. They represented organisations who either are or could be involved in commissioning, developing, implementing, and evaluating interventions that involve nature and health.

**Table 1.**
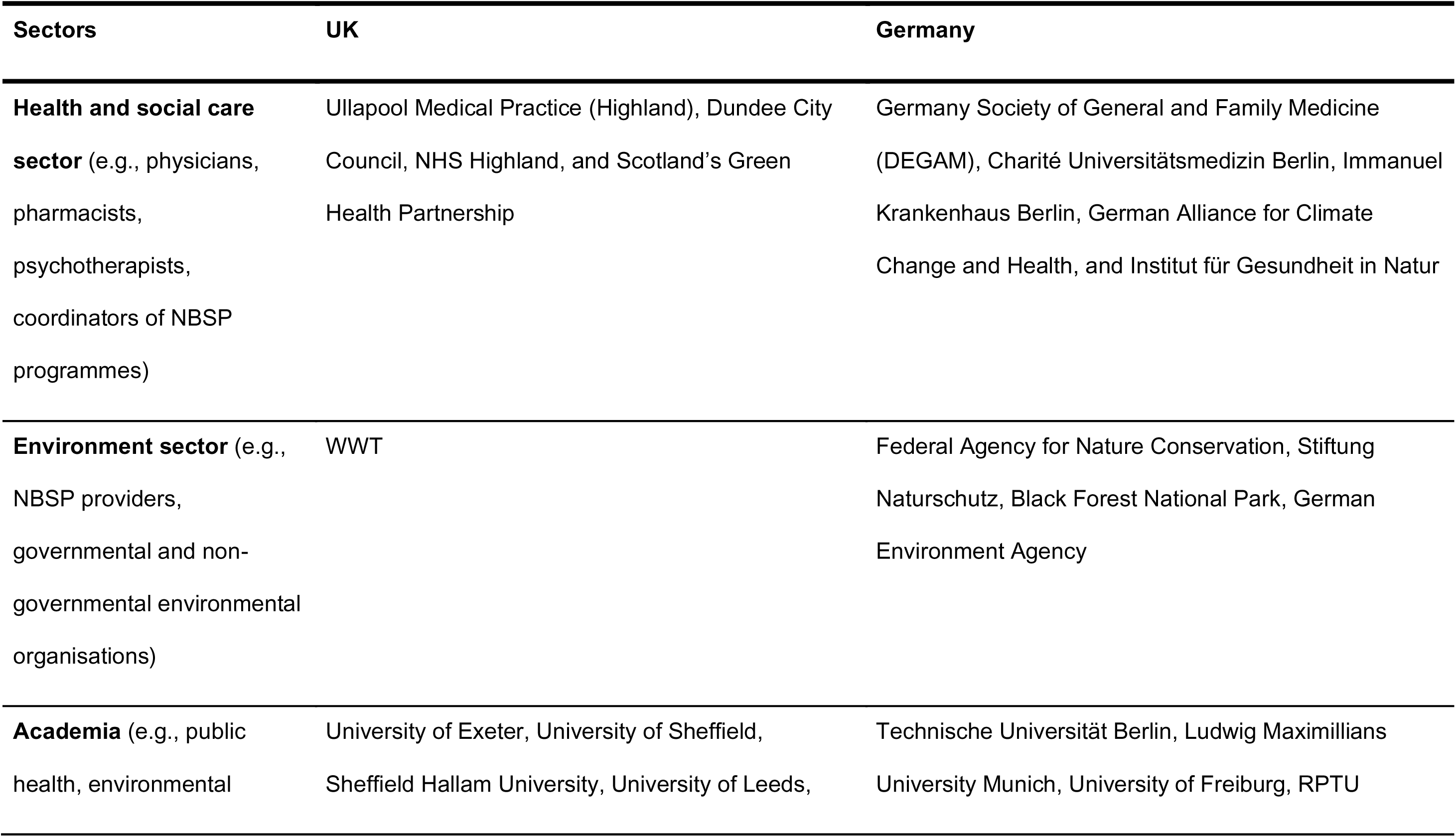
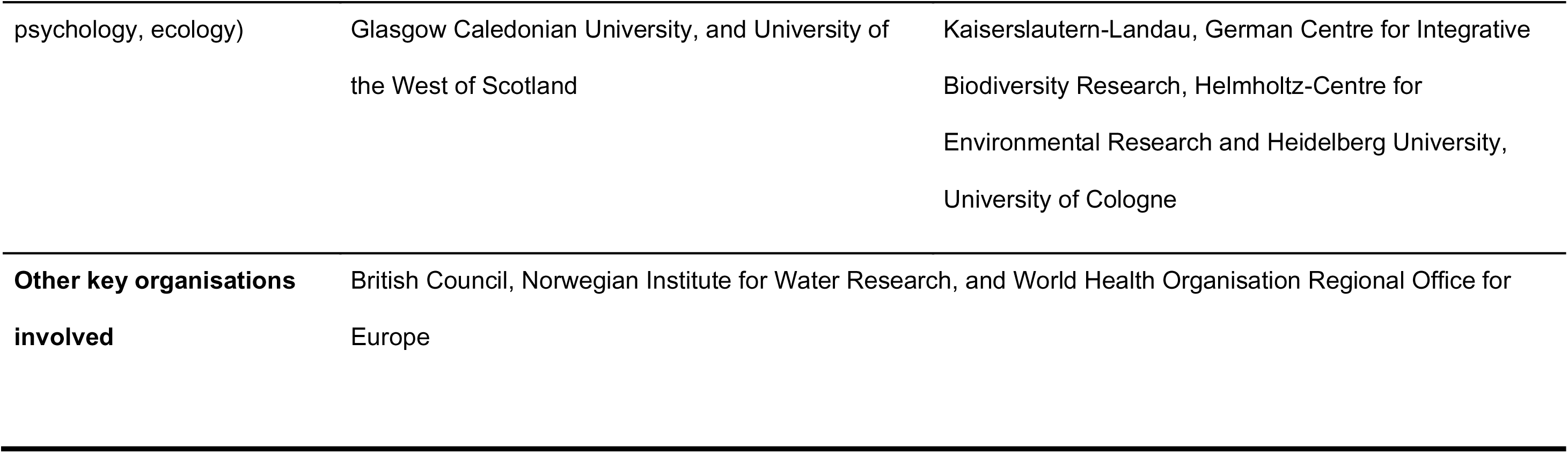
Organisations represented in the expert workshop on nature-based social prescribing.

### Workshop procedure

We used participatory knowledge-sharing approaches which included a mix of presentations, panel discussions, and breakout sessions (i.e., World Cafés). The workshop was divided into four sessions: Evidence and Current Practice; Environmental and Sustainability; Health and Wellbeing; and Policy and Financing (see Supplementary Material 1 for the detailed workshop programme). Presentations were intended to share best practice and create a mutual understanding of NBSP, thereby providing context for the panel discussions and World Cafés.

Between the sessions, we facilitated three World Cafés, each based on one of our aims (as stated above, for more detail see Supplementary Material 1), to allow in-depth discussion and knowledge exchange in smaller groups (54,55). In each World Café session, participants were divided into four sub-groups with assigned moderators and documenters. Each sub-group then focused on one of four topic areas: 1) existing and required evidence to capture the effectiveness of NBSP; 2) existing and required policies to implement NBSP; 3) existing and potential NBSP interventions; and 4) supporting and hindering factors of the healthcare system for NBSP. Each sub-group discussed their initial topic area for approximately 20 minutes before all except the moderator and documenter moved to the next topic area. Sub-groups were composed differently for each of the three World Cafés to facilitate diversity in the discussions. The discussions and ideas raised in the World Cafes were used to formulate recommendations for implementing pragmatic NBSP programmes in different contexts. By pragmatic, we mean the development of NBSP by building on current practice and working within existing parameters, for example, existing policy and healthcare system structures. Results of each discussion were captured on posters (Supplementary Material 2).

To allow for an open discussion, we followed the Chatham House Rules in all workshop discussions, which states that: *“the participants are free to use the information received, but neither the identity nor affiliation of the speaker(s), nor that of any other participant, may be revealed*” (56). This also enabled us to effectively “*bring people together, break down barriers, generate ideas, and agree solutions*” (56).

### Data analysis

We analysed and synthesised discussions from each World Café using (inductive) coding with thematic analysis (57). Outputs from each of the three World Cafés were initially analysed by one team member (SdB, JA, CM), then double-checked by a different team member (SdB, JA, CM). The analysis involved reading the notes and discussion from the World Café (information captured on posters in the workshop was transferred to an online interactive whiteboard for collaborative working) to identify descriptive themes (codes) from the workshop data. Three team members (SdB, JA, CM) then generated overarching (analytical) themes, in group discussions, by identifying and grouping similar descriptive themes occurring in all World Cafés. The themes and the codes were visualised using Google Jamboards (Supplementary Material 3).

Finally, to investigate more specifically key strengths, weaknesses, opportunities, and threats regarding the development and implementation of NBSP, we categorised Strengths, Weaknesses, Opportunities, and Threats (SWOT matrix) (58) within the overarching themes. The results of the thematic and SWOT analyses were presented to all workshop participants in an initial paper draft, giving everyone the opportunity to provide feedback on the synthesis, validate the findings, and ensure the accuracy and relevance of identified themes and their categorisation in the SWOT matrix.

## Results and Discussion

We identified five overarching themes from the World Café outputs (see Supplementary Material 3 for detailed results):

1. *Capacity building*, which relates to the ability of NBSP to deliver benefits to society, and the need for the development of the workforce to deliver NBSP;
2. *Universal accessibility*, which is concerned with the design and delivery of NBSP suitable for groups with different social and demographic characteristics;
3. *Embedded and integrated networks and collaborations*, which discusses how cross-sectoral networks at different levels are needed to support NBSP;
4. *Standardised implementation and evaluation*, which details the need for the development of typical (but adaptable) processes and procedures for delivering and evaluating NBSP; and
5. *Sustainability of the intervention*, which considers factors such as funding which are required for the continued delivery of NBSP.

Points raised by workshop participants were further categorised using a SWOT analysis (Figure 1, Table 2), to provide specific recommendations for the delivery of pragmatic NBSP programmes in different contexts within each of the five themes.

**Figure 1.**
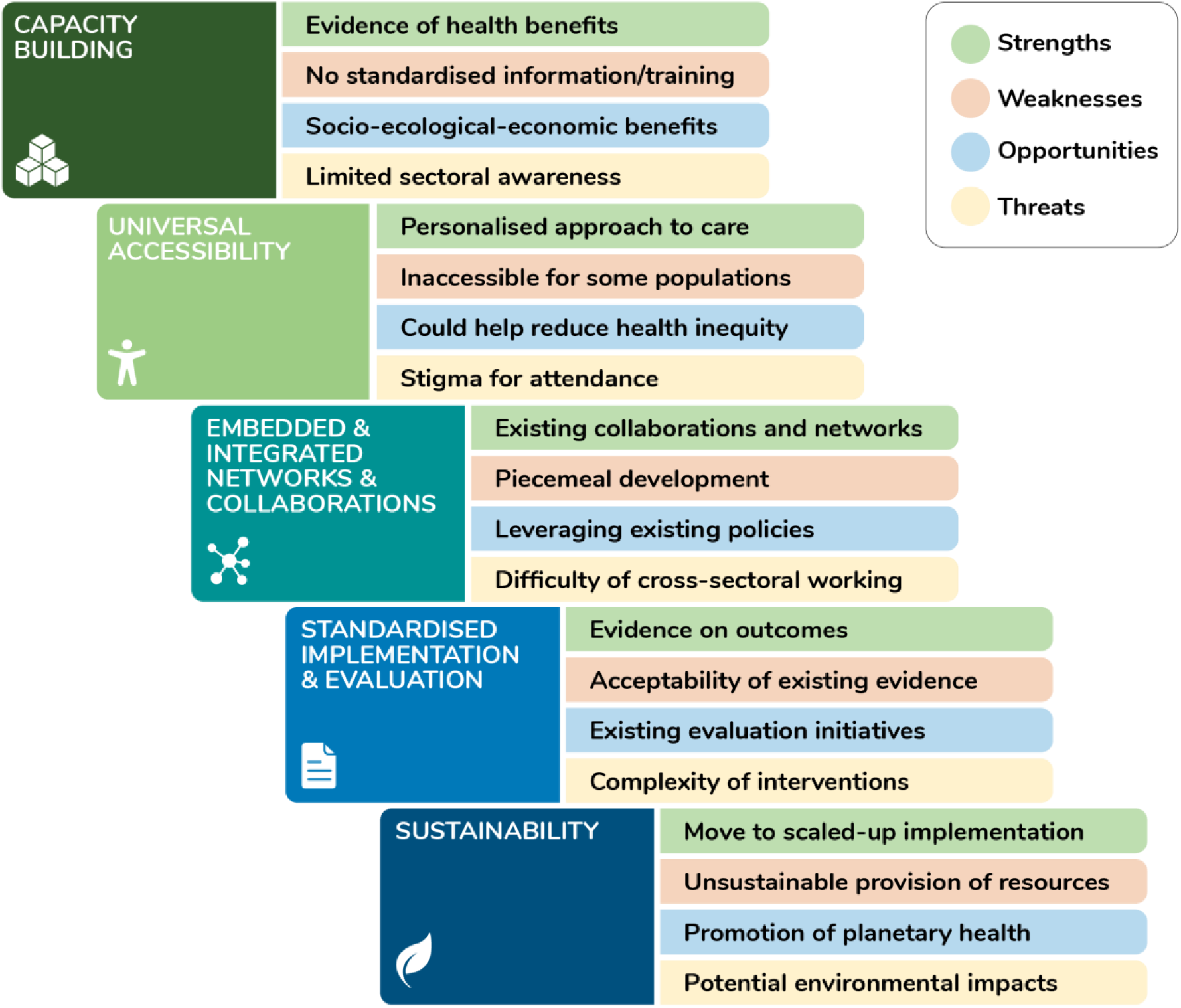
Summary of strengths, weaknesses, opportunities, and threats of Nature-Based Social Prescribing programmes categorised within the overarching themes.

**Table 2.** Analysis of strengths, weaknesses, opportunities, and threats related to implementation of NBSP in Germany and the UK identified during the expert workshop, organised by overarching theme.

| Overarching themes | Strengths | Weaknesses | Opportunities | Threats |
| --- | --- | --- | --- | --- |
| **Capacity building** – the ability of NBSP to provide benefits and need for development of the workforce to deliver NBSP | • Evidence that NBSP leads to health and social benefits | • Poor communication of scientific evidence to decision makers and healthcare providers | • NBSP has the potential to provide additional health and economic (e.g., healthcare costs) benefits | • Lack of understanding from potential service users (i.e., desire for pharmaceuticals, a culture of ‘pill for every ill’) |
|  |  | • No standardised information/training for those delivering NBSP<br>• Limited national/international definitions and models of NBSP | • NBSP could improve nature connection, which is associated with increased wellbeing and pro-environmental behaviours | • Limited understanding of links between nature and health, and planetary health (i.e., not yet included in school or medical school curriculum) |
| Universal accessibility – design and delivery of NBSP for diverse groups | • NBSP can target the public or specific groups with clinical needs | • There is no universal coverage (e.g., through statutory health insurance) | • NBSP programmes can be adapted and offer a variety of activities to suit a range of needs and contexts | • Stigma or prejudices associated with attending NBSP |
|  | • NBSP can be accessed through a range of routes (e.g., pharmacies, community settings); and modes (e.g., online or through the phone as well as in person) | • Programmes are not accessible to all (e.g., age, culture, region, socio-economic status, mobility status) | • NBSP could lead to increased exposure to natural spaces for participants | • Provision of natural spaces is not yet fully integrated into development and planning (i.e., in urban or medical settings) |
|  | • Variety of activities can be included in NBSP programmes so that individual preferences and |  | • With special targeting, NBSP could decrease health inequity as disadvantaged groups generally benefit more from |  |
|  | needs can be addressed |  | nature exposure |  |
| **Embedded and integrated networks and collaborations**<br>– development of cross-sectoral networks at different levels to support NBSP | Existing collaborations and partnerships between health, environment, and third sector organisations | Piecemeal/non-systematic approach to setting up NBSP<br>Lack of co-design with community/potential users | Policies that support NBSP (e.g., health in all policies, social determinants of health, community wealth)<br>Create job opportunities in health and environment sectors (e.g., link workers or equivalents, park rangers, surfing coach) | Current focus on treatment not prevention<br>More responsibilities and time constraints for current healthcare workers (e.g., when GPs must do link worker tasks)<br>Difficulties of trans-agency/-ministerial/-departmental/-sectoral collaborations to facilitate flows of funding<br>Unrealistic expectations of |
|  |  |  |  | GPs, services users, and other relevant actors |
| **Standardised implementation and evaluation**<br>– processes and procedures for delivering and evaluating NBSP | • Qualitative and case study evidence available | • Lack of scientific evidence that is acceptable to decision makers (e.g., quantitative from RCTs, meta-analyses, systematic reviews)<br>• No best practice guidelines or standards for implementation | • Ongoing monitoring would allow corrections to help service users | • Lack of acceptance of different forms of evidence (e.g., qualitative, pragmatic, and transdisciplinary evaluations, not only ‘standard’ controlled trials) |
|  |  | • No or limited implementation of standards for evidence collection or evaluation framework | • Ongoing monitoring would provide observational evidence for evaluation | • Complexity and diversity of outcomes and interventions |
|  |  | Limited resources to collect data or analyse existing data |  |  |
| **Sustainability – factors required for the continued delivery of NBSP** | Existing services (e.g. health promoting spas or curative health resort programmes in Germany; pilot blue and green social prescribing programmes in the UK) can be adapted to be NBSP and could be scaled up for national implementation | Funding is not long-term<br>Schemes are not sustainable (i.e., need time-bound exit strategies)<br>Lack of coordination in responsibilities and funding across sectors (i.e., health vs. environment) | Promotes planetary health (e.g., by increasing nature connectedness, and by reducing pharmaceutical use and/or the number of visits to the doctor)<br>Schemes to incentivise implementation and service users (e.g., reduced tariffs, which could also support interventions financially) | Potential for environmental degradation and biodiversity loss due to overuse |

**Note.** NBSP = nature based social prescribing. RCTs = randomised controlled trials. GP = general physician.

Below, we present a combined results and discussion section so that examples of current practice and research shared by workshop participants can be understood in context. This section is structured by the five themes identified.

### Capacity building

Building capacity for NBSP within the healthcare system is important to introduce and develop NBSP programmes that support and improve health and wellbeing at a population level. Whether resulting directly from participation in NBSP or as an indirect result of these programmes, evidence suggests that participation in NBSP can improve both physical and mental health. A recent meta-analysis showed reductions in blood pressure as well as anxiety and depression scores among service users ((59) see also (60) for related findings), while another recent review suggested that NBSP can improve social connectedness and mental wellbeing (18). Regarding health service delivery, NBSP may lead to reductions in the use of health services (e.g., pharmaceutical use) with associated economic and environmental benefits. This is supported by previous research indicating, for example, that social prescribing results in fewer visits to the GP and has a positive social return on investment (61).

Challenges discussed by participants relating to current capacity to deliver NBSP were similar to those recently identified for social prescribing programmes in integrated care (62). Capacity building is needed within three main areas. Firstly, there is limited information and training for professionals who might deliver NBSP. Education and training are necessary to equip professionals with the knowledge, skills, and competencies to deliver quality care. Depending on the NBSP pathway used, the professionals involved may include primary and secondary care practitioners (e.g., nurses and GPs), and community workers (e.g., nature-based activity managers, volunteers delivering activities). Participants discussed how some of these (e.g., link workers and community connectors) may not necessarily need a medical qualification, but they should be trained in relevant skills and competencies (e.g., in behaviour change methods, motivational interviewing, health assessment, action planning, safeguarding individuals with mental health conditions, coping strategies for themselves), as their role is crucial to the success of the referral, with many service users opening up more to them than to other healthcare professionals. This could be achieved through formal education programmes, specialised and accredited training courses, and/or continuous professional development to stay updated with the latest developments. This is likely to be context-specific as different countries have different professional training requirements. In the UK, for example, training might be adapted from cognitive behavioural therapy, as these courses have previously been developed to train a non-professional workforce (63).

Secondly, there is need for workforce expansion. Increasing NBSP provision means more professionals will be necessary to meet the growing demand for services (e.g., currently there is unmet demand for NBSP in the UK) (17). This may involve the creation of new professional roles. Whilst in the UK, link workers connect service users with appropriate nature-based activities, this role could be taken by other staff in the healthcare system in other countries. In Germany, it might be taken by GPs or social workers, as was discussed by workshop participants. Other countries operate with equivalents to link workers, such as the Netherlands with wellbeing coaches, Singapore with wellbeing coordinators, and the Philippines with village health workers and nutrition scholars (12,64,65). Participants also discussed strategies to recruit and retain staff (e.g., the provision of appropriate training and support). In the UK, there is currently high turnover in some roles because, for example, staff feel unsupported when service users share difficult stories (66).

To address the needs of an expanding workforce for knowledge, it may help to foster a broader understanding of the benefits of nature through education on planetary and public health, including the links between the environment and health, and giving opportunities to experience nature (e.g., through school and medical university curricula). In Germany, planetary health is not mandatory in medical training and education. However, education options are growing: for example, the German NGO “KLUG” has introduced online courses that address these issues within their own “Planetary Health Academy” (67). Also, the Chair of Public Health and Health Services Research has developed an online planetary health course for students for the “Virtual University of Bavaria” (68). Both programmes were funded by the German Government via the German Environment Agency. There is the potential for further development in this area; for example, there are ongoing clinical studies investigating the efficacy of nature and forest therapy at different German institutions, which could contribute materials for dissemination. While planetary health and sustainability are taught by many medical schools in the UK, teaching does not necessarily reflect current knowledge in these fields and is not mandatory (69).

Thirdly, along with the development of the workforce, to enable the presence of staff such as link workers, there is a need for the development of infrastructure and processes to enable the provision of NBSP in all health and social care facilities. For example, the UK is ensuring that everyone can access social prescribing through their GP by employing a sufficient number of link workers (70). However, expansion of NBSP will require these professionals being aware of available NBHI in their communities as insufficient communication across sectors can be a barrier (see also *Embedded and integrated networks and collaborations*). Currently, policymakers tend to give NBSP and other public health prevention programmes insufficient priority as their potential health-related benefits and co-benefits (i.e., climate change mitigation and adaptation, social cohesion, and nature conservation) have not yet been communicated effectively or are difficult to communicate due to the long timeframes until effects are visible. Developing good communication campaigns about nature’s health benefits could facilitate this acceptance (e.g., by land owners), and facilitate capacity building and the development of infrastructure and processes.

Related to finding effective communication strategies for policy makers, knowledge dissemination could also lead to increased service user acceptance, as they may have limited trust in NBSP due to a lack of understanding of the benefits of social prescribing (71) and NBSP (17). In this context, potential stigma for attending NBSP was discussed. Expectations and knowledge of service users are important as the beneficial effects of social prescribing programmes can be hindered when the aims, expectations, and/or interest of service users do not meet those of link workers or other people involved in the programme (72). The use of social marketing strategies (e.g., with buzz words) might help raise awareness amongst key audiences (cf. (73,74)).

### Universal accessibility

The benefits of nature exposure are often greater for deprived groups (75), meaning NBSP offers opportunities to address health inequalities. Whilst these benefits are less likely to be utilised for health promotion in countries without formalised NBSP practice, participants identified challenges relating to universal access that exist both in countries with existing structures for the implementation or regulation of NBSP, and those without (12).

Equity is a key issue, meaning that NBSP should be universally accessible to the population, particularly vulnerable groups. This would require equity to be addressed in each phase of implementation of NBSP activities, and consideration of the provision of prescriptions and referrals both at the individual and systems level. When developing an NBSP programme, a Health Equity Impact Assessment (76) might be useful in order to identify how the programme could reduce health inequalities and consider the perspective of individuals accessing NBSP and their specific needs. Programmes need to be inclusive of different individuals and population groups (e.g., people of different ages, cultural backgrounds, or with time and accessibility constraints). Individual characteristics and values associated with connecting to nature are important in understanding the many ways in which people are motivated to interact with the environment to successfully promote health and pro-environmental behaviours (31,77,78). Recognising that people’s relationships with and responsibilities for nature are diverse (79) can ensure that NBSP activities are inclusive and ultimately effective. In the UK, the Green Social Prescribing evaluation report found that targeting NBSP for specific groups could help reduce health inequities (17). This could be achieved through a varied service provision, for example with activities that stimulate different senses or that provide fun for people who are not intrinsically motivated by nature. Practical factors are also integral to accessibility, such as providing transport or considering the format of the activity. Integrating safe green and blue spaces in urban planning and hospital design could help to facilitate access to nature-based activities for both general populations and those with specific needs. When physical access is not possible, experiencing nature indirectly (e.g., virtually) can also provide benefits (80,81). In Scotland, the COVID-19 pandemic facilitated the development of virtual nature-based activities such as virtual health walks (Marx & More, 2022). In Germany, NBHI are also being delivered online (e.g., the Re.Connect Programme provides eco-psycho-social transformation activities (82)). Virtual programmes could supplement direct nature contact NBSP programmes to allow accessibility for certain populations (e.g., people with mobility issues).

Universal access to healthcare programmes largely depends on the resources, particularly financial, of the healthcare system. Differences in financial resources and approaches between different healthcare systems (e.g., publicly-funded or insurance-based) might create difficulties and hinder the implementation of NBSP (see also (62) for social prescribing in integrated care). In England, the government supports access to social prescribing by employing link workers (70), whereas in Scotland, there is limited funding to employ link workers. In both countries, NBSP activity providers are reliant on charitable and donor organisations. In contrast to the UK, NBSP in Germany would be an addition to the existing and rather complicated interplay of medical and social care provision. In Germany, it is compulsory for citizens to be insured by a (statutory or private) health insurance company against illnesses, accidents, and other health issues. Different German statutory and private insurance policies tend to include prevention strategies, mainly oriented to the prevention of specific diseases (or risk factors), for example by funding certain sports classes. Even though the benefits of spending time in nature are promoted by some German insurance companies (e.g., their websites have recommendations on ‘micro-adventures in nature’ (83) and forest bathing (84)), health insurance policies do not usually cover nature-based prevention strategies. This may also explain why uptake of NBSP in Germany is slow as there is no national champion to advocate for its mainstreaming in the healthcare system yet. Providers of health and social services could work together to make the case for NBSP to stakeholders.

There are specific routes that could be used to enable broad access to NBSP in Germany and elsewhere, such as funding link workers (or health professionals with similar roles) through statutory health insurances instead of obtaining individual funding through social enterprises. If a link worker structure was developed, it would be critical for their work to be coordinated with local primary care physicians. Service users who are regularly seen by secondary care physicians like cardiologists or endocrinologist (i.e., specialists) could also benefit from NBSP by either having link workers in secondary and tertiary settings (specialised care settings, often in hospitals) or by sending service users back to their primary care practitioners. Expanding the type of professionals who make NBSP referrals (i.e., identifiers) to, for example, pharmacists or secondary health care professionals, could reach a wider range of people in both countries, as some groups might be more likely to visit a pharmacy or a community centre than their GP. In Germany, NBSP referrals could also be provided in government-funded health kiosks (‘Gesundheitskiosk’), which are aimed at offering low-threshold counselling to people in socially disadvantaged districts.

### Embedded and integrated networks and collaborations

Enabling universally accessible NBSP programmes requires support by established interdisciplinary networks and collaborations, integrated across multiple sectors (especially health and environment government agencies, academia, and third sector organisations). Whilst there are examples of partnerships between health, environment, and third sector organisations that support or deliver NBSP, a major barrier to embedding and scaling up these collaborations is currently the low priority given to public health promotion and prevention programmes by policymakers in general. In many countries, including Germany and the UK, pharmaceutical treatment prevails as the most common form of healthcare intervention (85). Moving from treatment to prevention would require a shift in mindset among stakeholders, needing a whole systems approach (e.g., planetary health) and the insight that nature conservation is a prerequisite for health. Internationally, policies that are based on the planetary health approach could support NBSP, including the WHO Global Strategy on Health, Environment and Climate Change, Montreal-Kunming Global Biodiversity Framework, IUCN Global Standard for Nature-based Solutions, and the COP26 Health Programme (85–88).

Further challenges relate to the dominance of siloed thinking (i.e., a focus on sectoral priorities), and a resulting lack of collaboration and flow of funding across sectors. In this workshop, for example, there was limited representation in some areas (e.g., health insurance companies and politicians) although we invited representatives. In the UK, despite a ‘health in all policies’ (HiAP) approach, cross-sectoral policy-making is still challenging (e.g., due to the focus on treatment rather than preventative interventions in the health sector (89)). Policymaking should support interagency and sectoral collaborations, with shared action plans and goals. This top-down approach would need to be complemented by a bottom-up approach, to allow meaningful engagement and community co-creation, and ensure that NBSP programmes would be the go-to option when they are fully integrated in the healthcare system.

Networks and collaborations need strategies for knowledge exchange and sharing best practice. In the UK, ‘champions’ advocate for the HiAP approach and include both local authority and healthcare staff (89). These champions could similarly support NBSP, both as a voice in the community, promoting NBSP as part of supporting service users to take a self-care approach to their health, and by gaining support from policymakers, who have the power to make institutional change to enable NBSP.

### Standardised implementation and evaluation

Despite the quantity of research on the links between nature contact and health in general, a lack of sufficient scientific evidence on NBSP currently hinders standardised implementation and evaluation. Randomised controlled trials and systematic reviews based on these are seen as the ‘gold standard’ in the medical sciences. Without them, it is difficult to convince healthcare professionals and policymakers to commission NBSP, as decision-makers from the health sector, such as the German Joint Federal Committee which specifies the services that are reimbursed by statutory health insurance funds, rely heavily on these study designs. However, randomised controlled trials are expensive and often difficult to conduct in the case of NBSP interventions, while interdisciplinary and mixed methods approaches might be more appropriate for evaluating NBSP programmes. Although evidence based on well-designed studies is needed, previous research indicates that health policymakers tend to make decisions based on other factors, such as personal contacts, timeliness, and research containing easy-to-understand policy recommendations ((90); see also (73)), so the quality or type of scientific evidence might not be as significant a barrier as expected. High-quality research is still important, but consideration should be given to suitable communication too (e.g., through personal contact and readable summaries (74)).

The complexity and diversity of NBSP programmes and the range of relevant outcome variables is challenging and so is providing and interpreting the evidence. Standardised implementation and evaluation of NBSP will generate reliable evidence. In the UK, the implementation of social prescribing began locally and existed for many years before some aspects (e.g., funding of link workers) were embedded in NHS policies. NBSP has developed similarly in Canada (12), while in other countries, such as Austria, implementation of NBSP has been top-down (12). Whichever approach is taken, knowledge of the local context is important. Standardised national guidelines for implementation should be developed and adapted for individual programmes using local knowledge. This information could be collected through assessments of local community and health needs (e.g., conducting SWOT analysis, health needs analysis, or mapping of existing local resources). In addition, standardised but flexible and adaptable monitoring and evaluation frameworks, tools, and performance indicators should be developed. Whilst there are data security issues and reluctance by service users to provide (health) data that prevent analyses of available data, routine data could be used to develop a wider evidence base for NBSP. Furthermore, there is the potential for citizen science approaches and service user evaluation schemes to further help data collection. Regular and transparent programme monitoring will lead to ongoing quality control and assurance and ensure the identification of best practice in specific contexts by making the data freely accessible. It would also provide data to evaluate the impact of NBSP for a given service user in the long-term (e.g., on healthcare utilisation). This in turn would feed into standardised guidelines on best practice, including consideration of the impact of NBSP on the environment, and a competency framework for link workers.

Making NBSP part of clinical or prescribing guidelines would have benefits for its mainstreaming and delivery in different health and social care settings as it could help establish a tiered approach to delivering NBSP with other therapies such as talking therapy and medications. Currently, there is no accepted evaluation framework that would allow appropriate comparison of the effectiveness of NBSP programmes in improving health outcomes with existing clinical interventions (i.e., pharmaceuticals). Clinical interventions are usually evaluated using a straightforward cost-effectiveness analysis, under the biomedical model of health. However, NBSP programmes are complex interventions, stemming from the social model of health, which are more appropriately evaluated by a holistic evaluation mechanism. In the UK, the UK Social Value Act (2012) (91), Procurement Reform (Scotland) Act (2014) (92), and Wellbeing of Future Generations (Wales) Act (2015) (93) require commissioning and delivery organisations to be accountable for and consider how their NBSP programmes improve social, cultural, environmental, and economic wellbeing. The UK ministry for public finance and economic policy recommends the use of Social Cost Benefit Analysis (SCBA) for the evaluation of public health interventions (94). SCBA can determine whether the benefits of a programme offset the costs and convert this into monetary terms (94). Methodologically, Social Return on Investment (SROI) evaluation, when incorporated into a co-designed and co-produced approach from the outset (95), can be used to calculate monetary values for a wide range of outcomes of NBSP programmes, whether these already have a financial value or not. SROI analysis produces a description of how a NBSP programme generates value for key stakeholders and provides a Social Value Ratio, indicating how much social value (in £) is generated for every £1 of investment. For every £1 invested in a preventative NBSP, the social values generated range from £4.90 to £9.30 along with enhancing social cohesion, improving levels of health, wealth, and wellbeing (50,95).

Ultimately, NBSP programmes should be evaluated depending on their aim, to either prevent illnesses or to treat them. NBSP, and social prescribing in general, can be complementary to clinical interventions, with each having standalone benefits, meaning they could provide multiple benefits if used together. In Germany, current research projects on NBHI are being conducted with the aim to determine their treatment efficacy for different diseases. This provides a preliminary step towards possible implementation.

### Sustainability of the intervention

Limited resources hinder sustainable implementation of NBSP in both Germany and the UK. This includes resources for the delivery of NBSP (e.g., staff, time, opportunities for co-production of new NBSP programmes, follow-up support for vulnerable service users) and for evaluation. Currently, the implementation and evaluation of NBSP programmes often depends on funding from the third sector. Research shows this is a real barrier to NBSP in the UK (17) and in general (45,62). In Germany, the challenge of nationally upscaling the implementation of social prescribing relates to the fragmentation of health and social care finances across different sectors (12).

Integration and mainstreaming of NBSP in existing systems were considered important for programme sustainability. In the initial phase of NBSP programme development, resources already available in the community need to be identified (e.g., asset-based community development in the UK, drawing on existing community assets was found to be important by the Green Social Prescribing evaluation report (17)). This knowledge allows development of a long-term action plan, supported by a theory of change (a description of how the NBSP programme will benefit service users, with specifics of inputs, activities, outcomes, impacts). In Germany, there are four major prevention themes (diet, stress/resource management, addiction, physical exercise) financed by health insurance funds (96), in which some NBHI are already implemented (e.g., Nordic Walking or Tai Chi outdoors to increase mobility). In addition, highly specialised German health resorts offer traditional treatment programmes (called ‘Kur’) for various diseases based on available natural remedies (e.g., thermal water/mud) and a holistic approach (e.g., climatotherapy, Kneipp therapy, balneotherapy). Whilst there are new initiatives to develop recreational and therapeutic forests across Germany, such as forest bathing initiatives that aim at promoting people’s wellbeing through mindful immersive experiences in the forest (97,98), German health insurance companies do not yet regularly fund forest therapy. The number of rehabilitation clinics that implement forest bathing or forest therapy into their clinical setting has increased in the last years and could be extended to develop a community-based approach to NBSP. In the UK, programmes tend to last between 6 and 12 weeks while German health prevention seminars usually last 8 weeks (and can be up to 12 weeks). However, the ideal length of a programme depends on service users’ health status, needs, and other characteristics. Exit strategies are needed to support those leaving NBSP in maintaining benefits received from the programme and continuing activities independent of the programme.

Ultimately, a long-term action plan for the sustainability of NBSP should back investment in initiatives for prevention rather than reactive interventions. This might include incentivising both service providers and service users in a country-specific and appropriate way. This could be done by, for example, extra payments, refunds, or reduced health insurance tariffs. Explicitly linking NBSP to the wider social determinants of health and highlighting its potential benefits to health, climate, biodiversity, and environment (e.g., by reducing pharmaceutical pollution) in relation to cross-sectoral policies and programmes, might help policymakers to prioritise their use of resources, including those from non-health sectors, so that NBSP can be funded long-term.

Another barrier to the sustainability of NBSP programmes may be the availability and quality of green and blue spaces (17). Collaborations between the health, environment, and urban planning sectors as well as landowners are needed to ensure the availability and high-quality of natural space for NBSP activities. Potential damage of natural spaces due to unsustainable use of nature in NBSP programmes could also be problematic, as indicated by high usage of green spaces during the COVID-19 pandemic (99,100) being associated with various forms of damage, such as off-trail walking, littering, and increased disturbance to wildlife (101,102). This emphasises the importance of fostering responsible behaviour regarding proper usage of natural spaces in NBSP programmes (e.g., explaining potential restrictions in certain areas to safeguard wildlife or fragile ecosystems), and the necessity for allocating consistent funding to preserve the ecological integrity and long-term sustainability of natural spaces amid heightened demand. It is noteworthy that human wellbeing (esp. positive affect and meaningfulness), environmental education, ecological restoration, and pro-nature behaviour can go hand in hand (e.g. (103–105)), and can therefore be considered together when planning NBSP programmes. This relates also to the evidence of nature contact leading to nature connectedness (106), which is associated with pro-environmental behaviour (107,108). When planned well, NBSP programmes can promote human health and wellbeing, as well as nature connectedness and pro-environmental actions, likely leading to environmental benefits in the long run.

### Suggestions for policy, practice, and research

In summary, while NBSP programmes are increasingly recognised as a beneficial health intervention, work is still needed to ensure their effectiveness at different phases of development and scales of implementation (e.g., in the community and at the national level). Future implementation of NBSP in different contexts should build on the strengths and opportunities offered by current NBSP practice while considering its weaknesses and related threats (Figure 1, Table 2). Based on our workshop findings, we identified three key areas to progress NBSP:

- **Policy** should establish and expand cross-sectoral networks and collaborations (especially spanning the health and environment sectors), and incorporate actors from research, policy, and practice. This will enable cross-sectoral cooperation, shared understanding, standardised approaches, and co-funding opportunities to create the base needed to support sustainable NBSP.
- **Practice** should advance the development of standardised implementation and competency frameworks. These will enable the adaptation, accessibility, and evaluation of NBSP in different countries and contexts.
- **Research** should expand the evidence base to include a wider range of study designs and outcomes (e.g., socio-economic and ecological benefits), which could be used to better communicate the benefits of NBSP to different audiences, as well as identify factors crucial for successful implementation (in general and in specific contexts).

## Conclusions

There is increasing global interest in NBSP as a health intervention which could have much wider societal and environmental benefits, including a contribution to tackling the multiple crises facing the planet (109). In addition to promoting the physical, mental, and social health of individuals, participation in NBSP programmes could benefit healthcare systems by reducing demands for health services and pharmaceutical use, with associated reductions in financial costs. NBSP programmes could also have environmental implications, with the potential to increase nature connection and pro-environmental behaviours (106,108). However, NBSP faces a number of challenges to its implementation in different settings. While research shows the importance of nature for health, data for NBSP are limited due to the complexity of interventions and outcomes. Moreover, the available evidence is often poorly communicated to service users, practitioners, and policymakers. Effective NBSP programmes require cross-sectoral collaboration, but competing priorities and governance structures hinder prioritising and funding of NBSP.

The characteristics of a pragmatic NBSP programme will depend on different factors on the macro- (e.g., environmental crises), meso- (e.g., governance structures, healthcare system, society and culture, population health), and micro- (e.g., individual health and wellbeing status, preference for nature-based activities) levels. Nevertheless, there are common aspects which should be considered to facilitate adaptation. Developing pragmatic NBSP programmes will require capacity building, for example by creating training for providers and structures for working with services users in different healthcare systems. The accessibility of NBSP needs consideration to ensure universal access and to address practical aspects such as transport to the locations of the activities. Networks and collaboration across sectors are an essential part of this process. While NBSP programmes need to be specific to the context in which they are situated, standardised and adaptable protocols for implementation and evaluation will ensure that all relevant points are considered when adapting and evaluating programmes. This will support effective and sustainable implementation of NBSP and thereby foster nature-based solutions for public health and wellbeing.

## Data Availability

All data generated or analysed during this study are included in this published article [and its supplementary information files].

## List of abbreviations

DEFRA: Department for Environment, Food and Rural Affairs
GP: general practitioner
HiAP: Health in All Policies
KLUG: German Alliance for Climate Change and Health
NBHI: nature-based health intervention
NBSP: nature-based social prescribing
NHS: National Health Service
SCBA: social cost benefit analysis
SROI: social return on investment
SWOT: Strengths, Weaknesses, Opportunities, and Threats
WWT: Wildfowl & Wetlands Trust

## Declarations

### Ethics approval and consent to participate

Not applicable.

### Consent for publication

Not applicable.

## Acknowledgements

The programme builds stronger, more inclusive, internationally connected higher education and TVET systems. We thank the British Council and the British Embassy for hosting the workshop in Berlin, particularly A. Kienberger (Head of Education) and K. Robinson (Science & Innovation Officer). We also thank our workshop participants, many of whom are co-authors of this paper, and those who contributed their knowledge and expertise (F. Mayer, A. Villegas, C. Glahe, J-S. Baxter, and M. Eichinger).

AB and RRYO acknowledge the support of the German Centre for Integrative Biodiversity Research (iDiv) funded by the German Research Foundation (DFG-FZT 118, 202548816). JA acknowledges the support of Glasgow Caledonian University and the Hydro Nation Scholars Programme funded by the Scottish Government. RSS acknowledges the support from the Eva Mayr-Stihl Foundation. PM is staff member of the WHO, but the authors alone are responsible for the views expressed in this publication, and they do not necessarily represent the views, decisions, or policies of the WHO or their organisations.

## Authors’ contributions (CRediT author statement)

**SdB, JA, and CM**: Conceptualization, Methodology, Investigation, Resources, Formal analysis, Writing - Original Draft, Writing - Review & Editing, Visualization, Project administration, Funding acquisition **AB, RRYO, and MM**: Conceptualization, Methodology, Investigation, Resources, Writing - Review & Editing, Funding acquisition, Supervision. **RSS and TMS**: Conceptualization, Investigation, Writing - Review & Editing. **SB, MBG, MD, CD, JCF, AHa, AHe, GI, CSK, KK, MN, SP, CQ, JPR, KR, WS, KW, and CWN**: Writing - Review & Editing.

